# Humoral immune responses against SARS-CoV-2 Spike variants after mRNA vaccination in solid organ transplant recipients

**DOI:** 10.1101/2022.05.13.22275056

**Authors:** Alexandra Tauzin, Guillaume Beaudoin-Bussières, Shang Yu Gong, Debashree Chatterjee, Gabrielle Gendron-Lepage, Catherine Bourassa, Guillaume Goyette, Normand Racine, Zineb Khrifi, Julie Turgeon, Cécile Tremblay, Valérie Martel-Laferrière, Daniel E. Kaufmann, Marc Cloutier, Renée Bazin, Ralf Duerr, Mélanie Dieudé, Marie-Josée Hébert, Andrés Finzi

**Author notes:** Correspondence (A.F.), (M.J.H.).

## Abstract

While SARS-CoV-2 mRNA vaccination has been shown to be safe and effective in the general population, immunocompromised solid organ transplant recipients (SOTR) were reported to have impaired immune responses after one or two doses of vaccine. In this study, we examined humoral responses induced after the second and the third dose of mRNA vaccine in different SOTR (kidney, liver, lung and heart). Compared to a cohort of SARS-CoV-2 naïve immunocompetent health care workers (HCW), the second dose induced weak humoral responses in SOTR, except for the liver recipients. The third dose boosted these responses but they did not reach the same level as in HCW. Interestingly, while the neutralizing activity against Delta and Omicron variants remained very low after the third dose, Fc-mediated effector functions in SOTR reached similar levels as in the HCW cohort. Whether these responses will suffice to protect SOTR from severe outcome remains to be determined.

## INTRODUCTION

The severe acute respiratory syndrome coronavirus 2 (SARS-CoV-2) is the etiologic agent of the coronavirus disease 2019 (COVID-19) responsible of the current pandemic. COVID-19 causes a plethora of symptoms with different degrees of severity (Sheikhi et al., 2020). In solid organ transplant recipients (SOTR), due to immunosuppressive treatments, SARS-CoV-2 infection leads to a high rate of severe COVID-19 (Danziger-Isakov et al., 2021; Pereira et al., 2020) and therefore vaccination is strongly recommended (AST, 2022; CST, 2022). The Pfizer/BioNTech BNT162b2 and Moderna mRNA-1273 mRNA vaccines have shown a remarkable efficacy in the general population, particularly against severe outcomes (Baden et al., 2021; Polack et al., 2020). However, in SOTR, immune responses induced by vaccination are generally reduced (Kumar et al., 2011; Stucchi et al., 2018) and recent studies have shown that SOTR have impaired humoral responses after two doses of the SARS-CoV-2 mRNA vaccine (Caillard et al., 2021; Miele et al., 2021; Rabinowich et al., 2021; Rincon-Arevalo et al., 2021; Stumpf et al., 2021).

Moreover, SARS-CoV-2 is constantly evolving, and the Wuhan original strain has now been replaced by several variants. Among current circulating strains, the Delta and Omicron variants of concern (VOCs) have accumulated numerous mutations in their genome, and notably in the Spike (S) glycoprotein (Kumar et al., 2022). Because of the mutations, these VOCs are transmitted more efficiently than the original Wuhan strain and less well controlled by vaccination (Kumar et al., 2022; Lauring et al., 2022; Tseng et al., 2022). However, the administration of a third dose of mRNA vaccine (boost) leads to strong humoral responses and protects from severe outcome caused by these VOCs in the general population (Ariën et al., 2022; Tauzin et al., 2022a; Yoon et al., 2022). However, humoral responses elicited by the third dose on populations with compromised immune responses, particularly SOTR, are less documented. Here, we evaluatedhumoral responses induced in different groups of SOTR (kidney, liver, lung and heart) after the second and third doses of the mRNA vaccine.

## RESULTS

We analyzed humoral immune responses in cohorts of 31 kidney, 11 liver, 14 lung and 8 heart organ transplant recipients after the second (median [range]: 26 days [20–54 days]) and third doses (median [range]: 35 days [19–68 days]) of SARS-CoV-2 mRNA vaccine. The SOTR received their first two doses with different interval regimen (median [range]: 36 days [25–112 days]) and their third dose around 4 months after the second dose (median [range]: 110 days [34–195 days]), according to the province of Quebec, Canada public health authority’s vaccination roll out guidelines for immunocompromised patients. Vaccine-elicited humoral responses in SOTR were compared to those measured in a cohort of SARS-CoV-2 naïve health care workers (HCW). HCW received their first two doses of mRNA vaccine with a 16-week extended interval (median [range]: 111 days [76–134 days]), and their third dose around seven months after the second dose (median [range]: 219 days [167-235 days]), according to the province of Quebec, Canada public health authority’s vaccination roll out guidelines for HCW. Several studies have shown that this extended interval regimen leads to strong humoral and cellular responses after the second dose, notably against VOCs (Chatterjee et al., 2022; Nayrac et al., 2021; Payne et al., 2021; Tauzin et al., 2022a). This allowed us to compare humoral responses obtained in SOTR versus humoral responses elicited by a long interval vaccination regimen. Basic demographic characteristics of the cohorts, immunosuppressive treatments of the SOTR and detailed vaccination time points are summarized in Table 1 and Figure 1A.

**Table 1.**
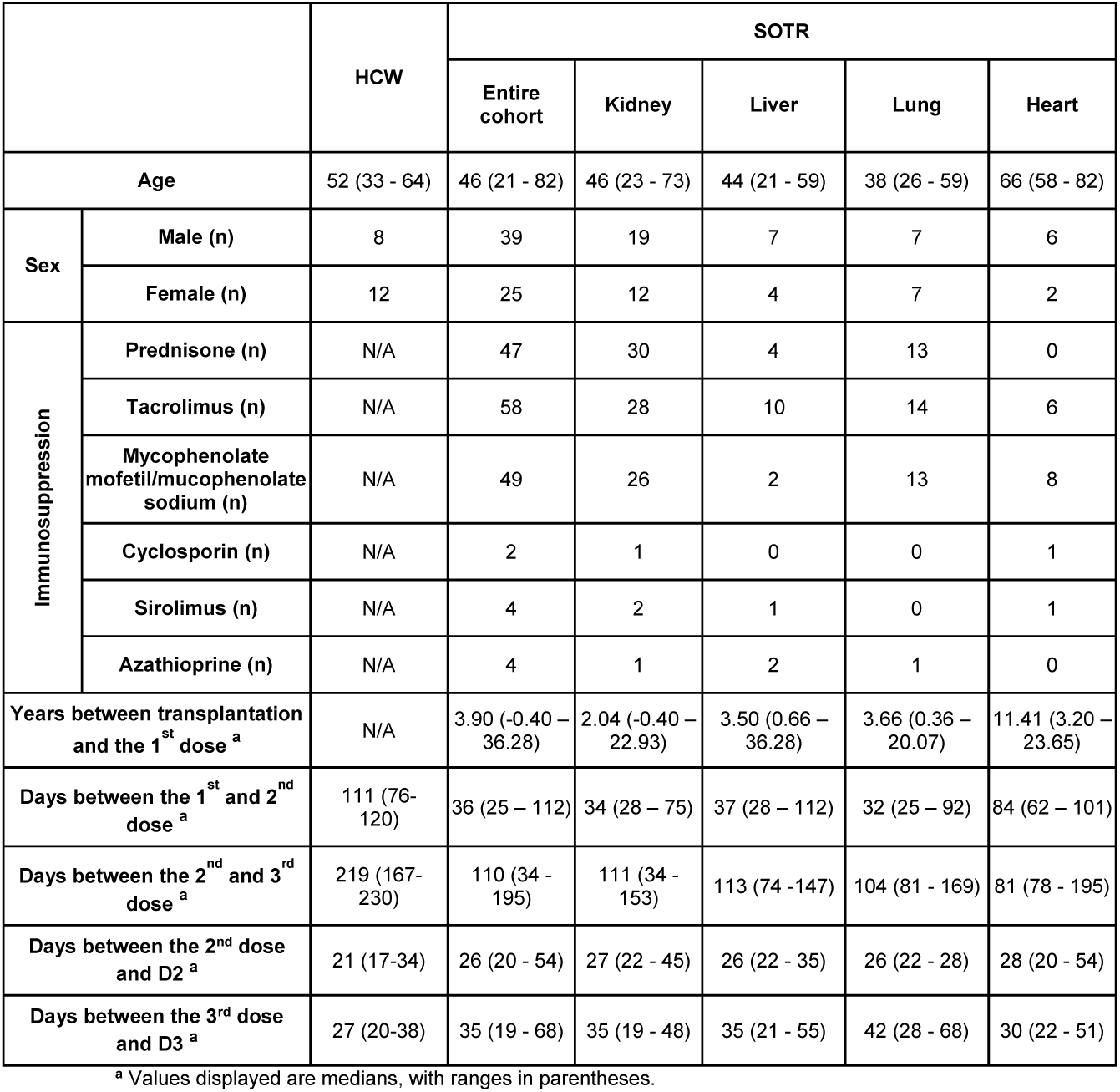
Characteristics of the SARS-CoV-2 vaccinated SOTR and HCW cohorts.

|  |  | HCW | SOTR |  |  |  |  |
| --- | --- | --- | --- | --- | --- | --- | --- |
|  |  |  | Entire cohort | Kidney | Liver | Lung | Heart |
| **Age** |  | 52 (33 - 64) | 46 (21 - 82) | 46 (23 - 73) | 44 (21 - 59) | 38 (26 - 59) | 66 (58 - 82) |
| **Sex** | **Male (n)** | 8 | 39 | 19 | 7 | 7 | 6 |
|  | **Female (n)** | 12 | 25 | 12 | 4 | 7 | 2 |
| **Immunosuppression** | **Prednisone (n)** | N/A | 47 | 30 | 4 | 13 | 0 |
|  | **Tacrolimus (n)** | N/A | 58 | 28 | 10 | 14 | 6 |
|  | **Mycophenolate mofetil/mucophenolate sodium (n)** | N/A | 49 | 26 | 2 | 13 | 8 |
|  | **Cyclosporin (n)** | N/A | 2 | 1 | 0 | 0 | 1 |
|  | **Sirolimus (n)** | N/A | 4 | 2 | 1 | 0 | 1 |
|  | **Azathioprine (n)** | N/A | 4 | 1 | 2 | 1 | 0 |
| **Years between transplantation and the 1<sup>st</sup> dose <sup>a</sup>** |  | N/A | 3.90 (-0.40 – 36.28) | 2.04 (-0.40 – 22.93) | 3.50 (0.66 – 36.28) | 3.66 (0.36 – 20.07) | 11.41 (3.20 – 23.65) |
| **Days between the 1<sup>st</sup> and 2<sup>nd</sup> dose <sup>a</sup>** |  | 111 (76-120) | 36 (25 – 112) | 34 (28 – 75) | 37 (28 – 112) | 32 (25 – 92) | 84 (62 – 101) |
| **Days between the 2<sup>nd</sup> and 3<sup>rd</sup> dose <sup>a</sup>** |  | 219 (167-230) | 110 (34 - 195) | 111 (34 - 153) | 113 (74 -147) | 104 (81 - 169) | 81 (78 - 195) |
| **Days between the 2<sup>nd</sup> dose and D2 <sup>a</sup>** |  | 21 (17-34) | 26 (20 - 54) | 27 (22 - 45) | 26 (22 - 35) | 26 (22 - 28) | 28 (20 - 54) |
| **Days between the 3<sup>rd</sup> dose and D3 <sup>a</sup>** |  | 27 (20-38) | 35 (19 - 68) | 35 (19 - 48) | 35 (21 - 55) | 42 (28 - 68) | 30 (22 - 51) |

<sup>a</sup> Values displayed are medians, with ranges in parentheses.

**Figure 1.**
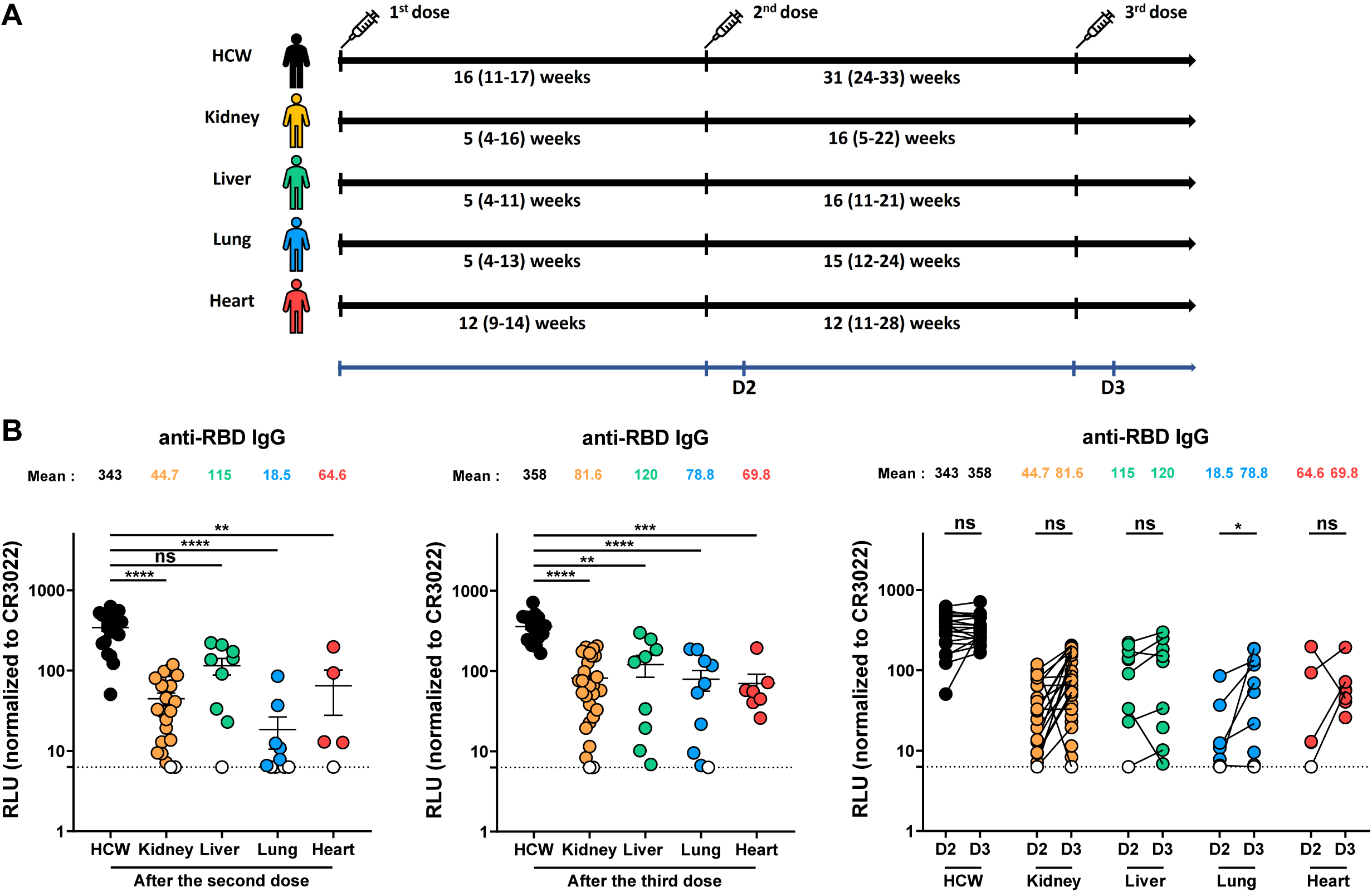
Elicitation of RBD-specific antibodies in SOTR and HCW after mRNA vaccination. (**A**) SARS-CoV-2 vaccine cohorts’ design. (**B**) Indirect ELISA was performed by incubating plasma samples from SOTR or HCW with recombinant SARS-CoV-2 RBD protein. Anti-RBD Ab binding was detected using HRP-conjugated anti-human IgG. Relative light unit (RLU) values obtained with BSA (negative control) were subtracted and further normalized to the signal obtained with the anti-RBD CR3022 mAb presents in each plate. **Left panel** shows the values obtained after the second dose, and **middle panel** after the third dose. **Right panel** shows the difference obtained between D2 (post second dose) and D3 (post third dose) for every group. Symbols represent biologically independent samples from SOTR and HCW. Lines connect data from the same donor. Undetectable measures are represented as white symbols, and limits of detection are plotted. Error bars indicate means ± SEM. (* p < 0.05; ** p < 0.01; *** p < 0.001; **** p < 0.0001; ns, non-significant).

### Elicitation of SARS-CoV-2 antibodies against the receptor-binding domain of the Spike

We first measured anti-receptor-binding domain (RBD) IgG levels induced after the second and the third doses of the mRNA vaccine using a previously reported ELISA assay (Anand et al., 2021; Beaudoin-Bussières et al., 2020; Prévost et al., 2020; Tauzin et al., 2021). After the second dose, all HCW presented high levels of RBD-specific IgG (Figure 1B). In contrast, in all groups of SOTR, the levels of anti-RBD antibodies (Abs) were, with the exception of liver recipients, significantly lower than in HCW. We also noted that in every SOTR group, some donors did not have anti-RBD IgG after the second dose of mRNA vaccine. Among SOTR, liver recipients had higher Ab levels than kidney, lung and heart recipients, in line with a generally lower immunosuppression regimen. For HCW, the third dose of the mRNA vaccine led to the same level of Abs as after the second dose, as recently described (Tauzin et al., 2022a). For SOTR, we observed a significant increase in the level of anti-RBD IgG in lung recipients and a trend for kidney and heart recipients. No increase in anti-RBD IgG level was observed for liver recipients. Of note, in all SOTR groups, anti-RBD levels remained significantly lower than in the HCW cohort even after the third dose, but most donors who did not have anti-RBD IgG after the second dose developed antibodies after the third dose, suggesting the initiation of an antibody response by repeated antigen exposure.

### Recognition of SARS-CoV-2 Spike variants and a common-cold human *Betacoronavirus*

We next evaluated the recognition of the SARS-CoV-2 full-length S after vaccination in SOTR and HCW by flow cytometry (Figure 2A). After the second vaccine dose, no significant differences were observed in the recognition of the D614G S by plasma from kidney and liver recipients and HCW. In contrast, lung and heart recipients recognized the D614G S less efficiently. The third dose increased the D614G S recognition for HCW, and we noted a slight increase for lung and heart recipients, for whom the recognition was very weak after the second dose. For kidney and liver recipients, the third dose did not improve the D614G S recognition (Figure 2A).

**Figure 2.**
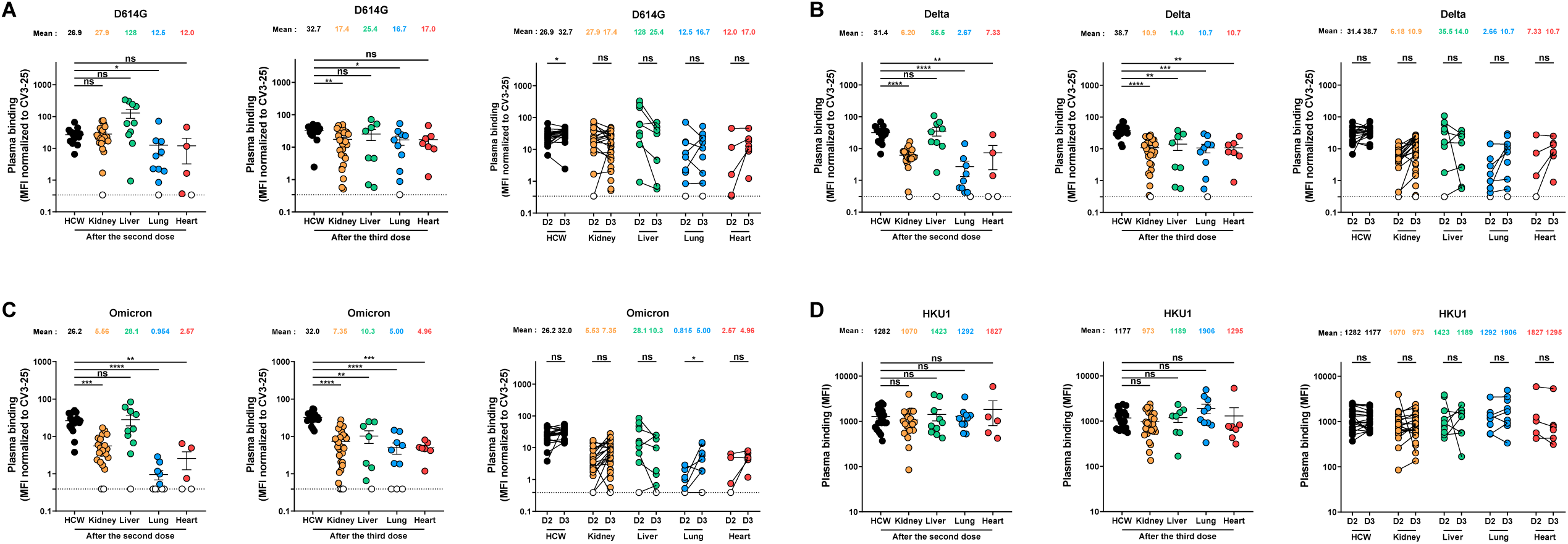
Binding of vaccine-elicited antibodies to SARS-CoV-2 Spike variants and the human HKU1 *Betacoronavirus* in SOTR and HCW after mRNA vaccination. 293T cells were transfected with the indicated full-length S from different SARS-CoV-2 variants and HCoV-HKU1 and stained with the CV3-25 Ab or with plasma from SOTR or HCW. The values represent the median fluorescence intensities (MFI) normalized by CV3-25 Ab binding (**A-C**) or the MFI (**D**). **Left panel** shows the values obtained after the second dose, and **middle panel** after the third dose. **Right panel** shows the differences obtained between D2 (post second dose) and D3 (post third dose) for every group. Symbols represent biologically independent samples from SOTR and HCW. Lines connect data from the same donor. Undetectable measures are represented as white symbols, and limits of detection are plotted. Error bars indicate means ± SEM. (* p < 0.05; ** p < 0.01; *** p < 0.001; **** p < 0.0001; ns, non-significant).

It has been well documented that Delta and Omicron VOCs are less efficiently recognized by Abs induced by vaccination, because of accumulated mutations in the S glycoproteins compared to the original Wuhan strain, used for the development of current mRNA SARS-CoV-2 vaccines (Chatterjee et al., 2022; Planas et al., 2021; Tauzin et al., 2022a). We measured the recognition of these VOCs S after mRNA vaccination in SOTR (Figure 2B-C). We did not see significant differences in the level of recognition of Delta and Omicron S between liver recipients and HCW after the second dose. The third dose led to a slight increase of the recognition of the VOCs S except for liver recipients, however it remained significantly lower than in HCW. When we compared S recognition between the SARS-CoV-2 variants (Figure S1), we observed that in HCW, because of strong humoral responses induced by the extended interval, no major differences in recognition were observed between D614G and VOCs S after the second and third doses of mRNA vaccine (Figure S1A). For SOTR, VOCs S were significantly less recognized than the D614G S, suggesting that vaccination in SOTR did not improve the breadth of S recognition, as observed in HCW (Figure S1B-E).

We also evaluated the recognition of the human HKU1 *Betacoronavirus* S glycoprotein (Figure 2D). HKU1 is an endemic coronavirus that causes common colds and is highly prevalent in the population (Chan et al., 2009; Rees et al., 2021). No significant differences between HCW and SOTR were observed after the second and the third doses of the vaccine, indicating that transplantation and associated immunosuppression regimens did not affect the level of circulating Abs elicited before vaccination.

### Functional activities of vaccine-elicited antibodies

We evaluated functional activities of vaccine-elicited Abs after the second and third doses of mRNA vaccine (Figure 3). We measured Fc-mediated effector functions using a well-described antibody-dependent cellular cytotoxicity (ADCC) assay (Anand et al., 2021; Beaudoin-Bussières et al., 2020, 2021; Ullah et al., 2021). Plasma from HCW presented robust ADCC activity after the second dose that was restored to the same level by the third dose (Figure 3A). The second dose elicited ADCC-mediating Abs in liver and heart recipients that reached similar levels of activity as in HCW. This is in contrast with significant lower ADCC activity elicited after the second dose in kidney and lung recipients. The boost led to a significant increase in ADCC activity in these donors. Importantly, the third dose elicited ADCC activity in all SOTR similar to the one observed in HCW.

**Figure 3.**
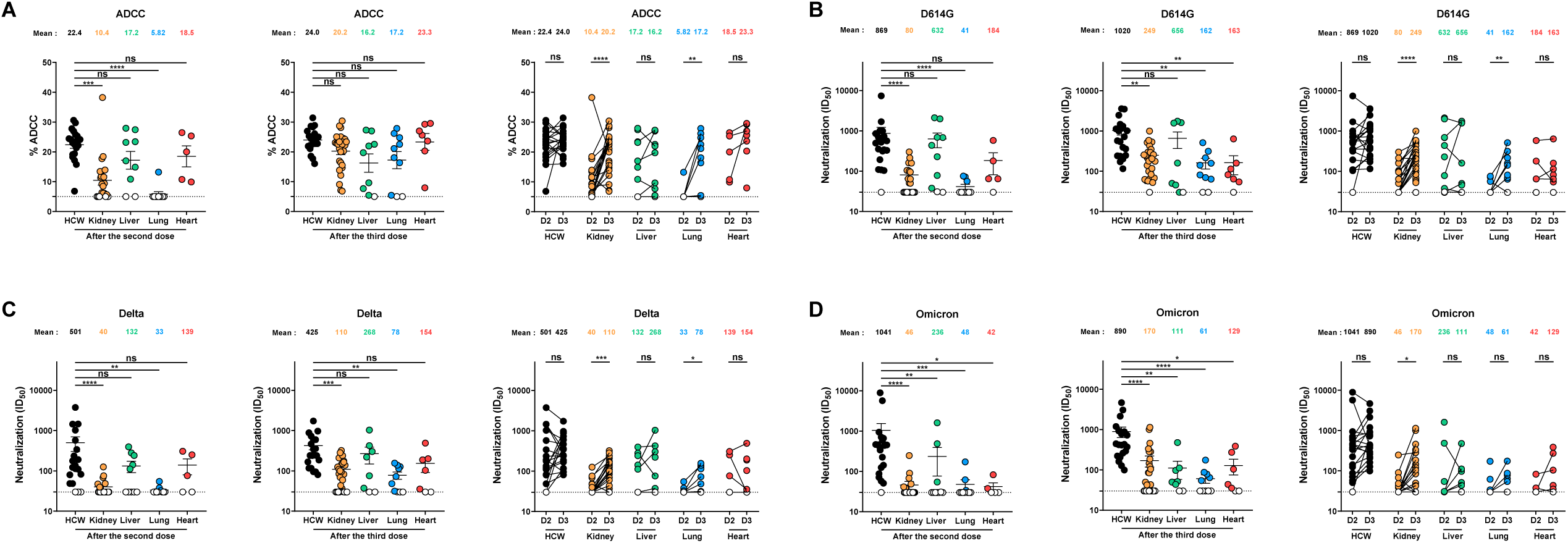
Fc-mediated effector functions and neutralization activities in SOTR and HCW after mRNA vaccination. (**A**) CEM.NKr parental cells were mixed at a 1:1 ratio with CEM.NKr-Spike cells and were used as target cells. PBMCs from uninfected donors were used as effector cells in a FACS-based ADCC assay. (**B-D**) Neutralizing activity was measured by incubating pseudoviruses bearing SARS-CoV-2 S glycoproteins ((**B**) D614G, (**C**) Delta and (**D**) Omicron), with serial dilutions of plasma for 1 h at 37°C before infecting 293T-ACE2 cells. Neutralization half maximal inhibitory serum dilution (ID_50_) values were determined using a normalized non-linear regression using GraphPad Prism software. **Left panel** shows the values obtained after the second dose, and **middle panel** after the third dose. **Right panel** shows the differences obtained between D2 (post second dose) and D3 (post third dose) for every group. Symbols represent biologically independent samples from SOTR and HCW. Lines connect data from the same donor. Undetectable measures are represented as white symbols, and limits of detection are plotted. Error bars indicate means ± SEM. (* p < 0.05; ** p < 0.01; *** p < 0.001; **** p < 0.0001; ns, non-significant).

We also measured the neutralizing activity of the vaccine-induced Abs, against pseudoviruses carrying SARS-CoV-2 S (Figure 3B-D). When assessing the neutralizing activity against the D614G S, we observed that the second dose elicited Abs with neutralizing activity in liver recipients (Figure 3B). In other SOTR, very low levels of neutralizing Abs were detected, especially in lung recipients. As observed for ADCC activity, the boost increased the neutralization activity in kidney and lung recipients. However, even after the third dose, SOTR did not reach the same levels of neutralizing Abs as in HCW.

We also measured the neutralizing activity against pseudoviruses carrying the Delta and Omicron Spikes (Figure 3C-D). In HCW, the second dose of mRNA vaccine administered with a 16-weeks interval, led to high levels of Abs able to neutralize these variants, as previously described (Chatterjee et al., 2022; Payne et al., 2021; Tauzin et al., 2022a). In contrast, SOTR elicited very low levels of neutralizing Abs against Delta and Omicron variants after the second dose and, although the boost led to a slight increase of the neutralization activity, this remained significantly lower than in HCW.

When comparing the neutralizing activity between the SARS-CoV-2 variants (Figure S2), Delta was less efficiently neutralized in HCW than D614G at the two different time points (Figure S2A). For kidney and liver recipients, Omicron and Delta VOCs were less neutralized than D614G (Figure S2B-C). For other SOTR, neutralization activity was too weak to measure significant differences (Figure S2D-E).

### Anti-RBD avidity of vaccine-elicited antibodies

We also used a surrogate assay for antibody maturation by measuring the avidity for the RBD of vaccine-elicited Abs, using a previously described assay (Björkman et al., 1999; Fialová et al., 2017; Tauzin et al., 2022a, 2022b, 2022c). Briefly, plasma samples were tested in parallel by ELISA with washing steps having or not having a chaotropic agent (8M urea), measuring respectively the level of IgG with high avidity for the RBD and the level of total anti-RBD IgG. The RBD-avidity index corresponds to the proportion of high avidity IgG among the total anti-RBD IgG (Figures 4), and provides an overall idea of antibody maturation (Björkman et al., 1999; Fialová et al., 2017; Tauzin et al., 2022a, 2022b, 2022c).

**Figure 4.**
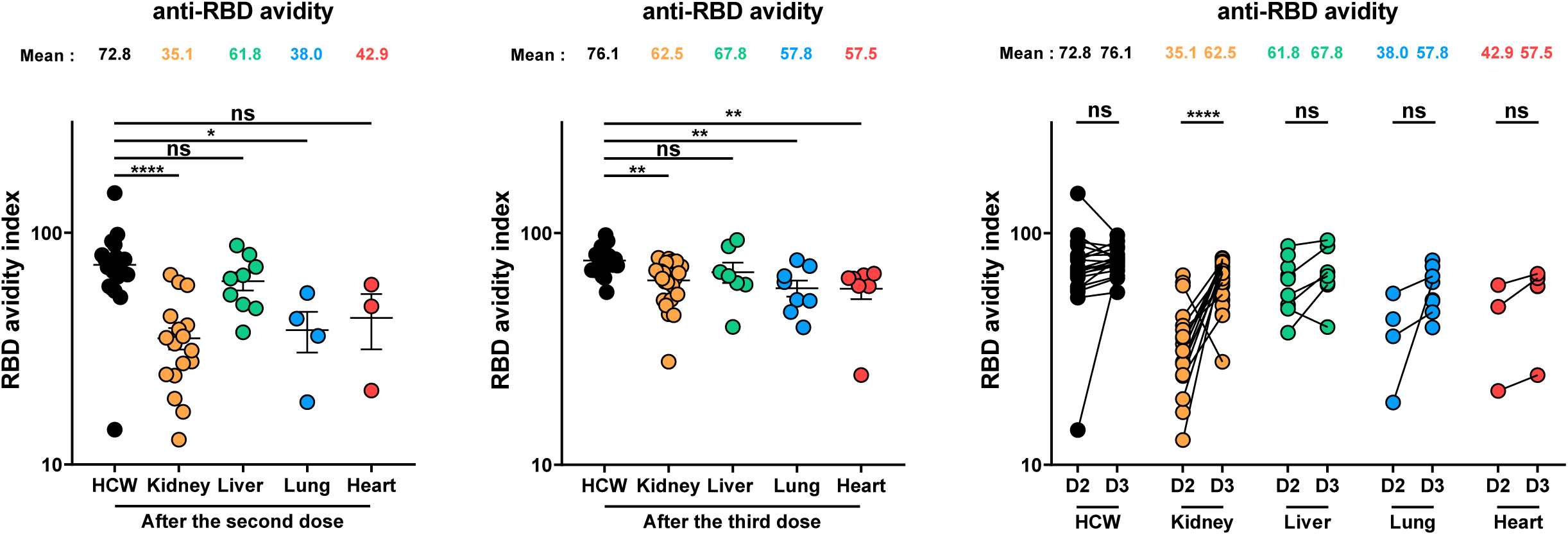
Anti-RBD avidity of vaccine-elicited antibodies in SOTR and HCW after mRNA vaccination. Indirect ELISA and stringent ELISA were performed by incubating plasma samples from SOTR and HCW with recombinant SARS-CoV-2 RBD protein. Anti-RBD Ab binding was detected using HRP-conjugated anti-human IgG. The RBD avidity index corresponded to the RLU value obtained for every plasma sample with the stringent (8M urea) ELISA divided by that obtained with the 0M urea ELISA. **Left panel** shows the values obtained after the second dose, and **middle panel** after the third dose. **Right panel** shows the differences obtained between D2 (post second dose) and D3 (post third dose) for every group. Symbols represent biologically independent samples from SOTR and HCW. Lines connect data from the same donor. Undetectable measures are represented as white symbols, and limits of detection are plotted. Error bars indicate means ± SEM. (* p < 0.05; ** p < 0.01; **** p < 0.0001; ns, non-significant).

In HCW, the second dose of the mRNA vaccine elicited IgG with high avidity, that was not further improved by the boost (Figure 4), as recently described (Tauzin et al., 2022a). In contrast, in SOTR who developed Abs able to recognize the RBD, the avidity was significantly lower than in HCW (Figure 1B and 4). The third dose of mRNA vaccine increased RBD avidity in SOTR but, with the exception of liver recipients, remained significantly lower than in HCW.

### Integrated analysis of vaccine responses elicited in solid organ transplant recipients

We evaluated the network of pairwise correlations among all studied immune variables on the HCW and the different SOTR groups (Figure 5). For HCW, we observed that after the second dose all immune variables tested were involved in a dense network of positive correlations. After the boost, we did not observe major differences in the network of correlations, suggesting that the third dose did not induce qualitatively different humoral responses in HCW. For lung and heart recipients, who received the strongest immunosuppressive regimens, we observed that all immune variables were very weakly interconnected after the second dose and the third dose did not strongly increase the network. Immune variables were slightly more interconnected for kidney recipients, which aligns with their lower immunosuppressive regimen compared to heart and lung recipients. For liver recipients, the network of correlation was less dense than in HCW after the second dose as observed for the other groups of SOTR. Interestingly, in this less immunosuppressed group of SOTR, the third dose of the mRNA vaccine induced a dense network of correlations, which was in a comparable range as in HCW.

**Figure 5.**
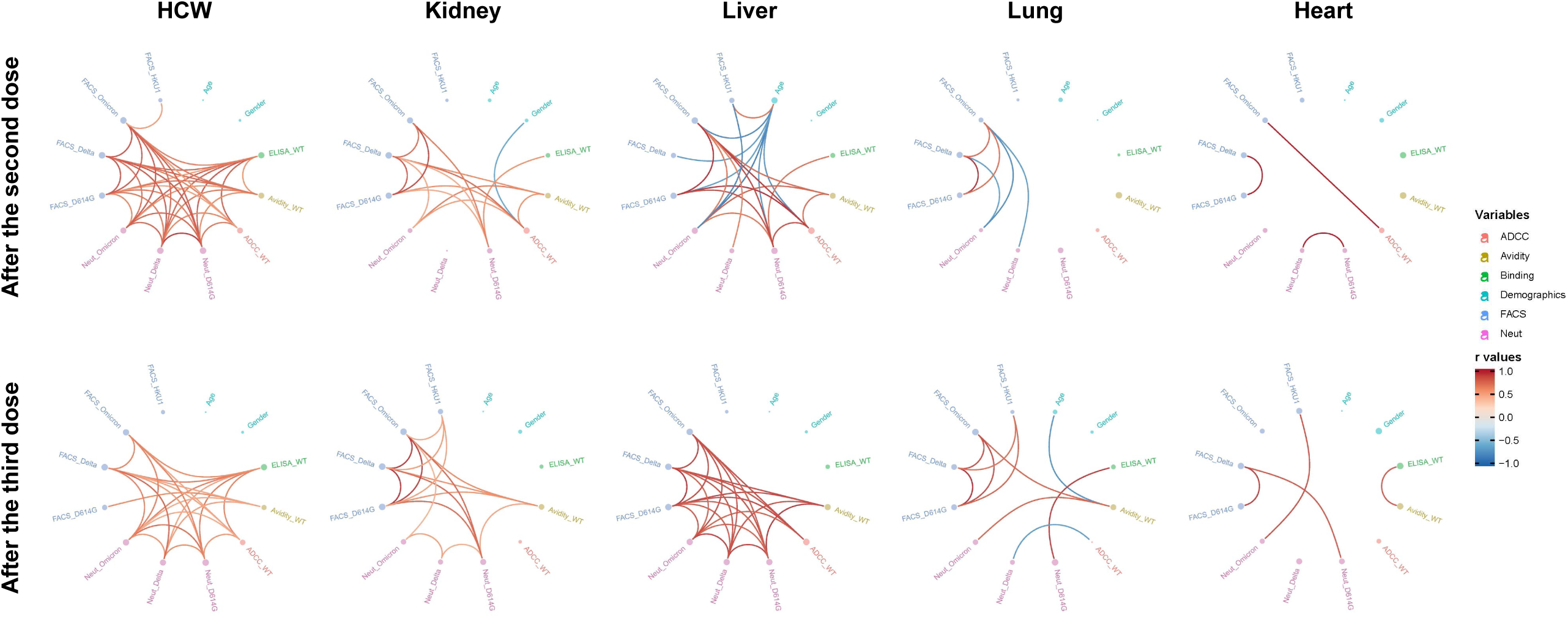
Mesh correlations of humoral response variables in SOTR and HCW after mRNA vaccination. Edge bundling correlation plots where red and blue edges represent positive and negative correlations between connected variables, respectively. Only significant correlations (p < 0.05, Spearman rank test) are displayed. Nodes are color coded based on the grouping of variables according to the legend. Node size corresponds to the degree of relatedness of correlations. Edge bundling plots are shown for correlation analyses using ten different datasets, i.e., HCW and kidney, liver, lung or heart transplant recipients after the second and third doses of mRNA vaccination.

## DISCUSSION

While a large part of the world population is vaccinated with two or three doses of SARS-CoV-2 vaccines, some population groups remain vulnerable to SARS-CoV-2 infection and most importantly to severe outcomes. Here we show that SOTR, known to respond less efficiently to vaccination due to their chronic immunosuppressive regimen (Kumar et al., 2011; Stucchi et al., 2018), elicited poor humoral responses after the second dose of SARS-CoV-2 mRNA vaccine, compared to HCW. The boost induced an increase of these responses, but they did not reach the same level as observed in HCW.

An important concern about the evolving pandemic is the frequent apparition of variants. It was previously shown that the 3-4 weeks standard interval of vaccination leads to weak neutralizing Abs against several VOCs in the general population (Chatterjee et al., 2022; Payne et al., 2021; Tauzin et al., 2022a, 2022c). However, administering a boost strongly enhances the breadth of neutralization activity against these variants (Nemet et al., 2022; Schmidt et al., 2022; Tauzin et al., 2022a). In SOTR, we did not observe a significant increase in the breadth of recognition and neutralization of these variants, suggesting an inability in Abs maturation in most of these individuals. This is supported by the poor anti-RBD avidity detected in these individuals, likely reflecting poor B cells maturation compared to HCW.

Interestingly, we observed that SOTR elicited Abs with ADCC activity comparable to HCW after the third dose of the mRNA vaccine. There is increasing evidence showing that Fc-mediated effector functions play an important role in the protection against severe outcomes of SARS-CoV-2 (Anand et al., 2021; Richardson et al., 2022; Tauzin et al., 2021). However, whether this will suffice to protect SOTR from severe outcomes caused by SARS-CoV-2 remains unknown.

We also noted some differences in humoral responses, depending on the transplanted organ. Notably, we observed that liver recipients had better humoral responses than other SOTR groups. These differences are probably due to the lower immunosuppressive regimens in liver recipients than in other SOTR groups. Further work is needed to understand the correlation between specific immunosuppressive regimens and vaccination outcome, taking into account the dose and type of immunosuppressive agents, and response to SARS-CoV-2 vaccines. This also highlights the importance of evaluating the different SOTR groups independently regarding the decisions on the follow-up of vaccinations that needs to be adapted to each SOTR group.

## Data Availability

All data produced in the present work are contained in the manuscript

## ACKNOWLEDGMENTS

The authors are grateful to the donors who participated in this study. The authors thank the CRCHUM BSL3 and Flow Cytometry Platforms for technical assistance. We thank Dr. M. Gordon Joyce (U.S. MHRP) for the monoclonal antibody CR3022. We also thank Demitra Yotis, Amani Mahroug, Yizou Zhao and Annie Karakeussian Rimbaud for their contribution on the transplant cohort. The graphical abstract was created using BioRender.com. This work was supported by le Ministère de l’Économie et de l’Innovation du Québec, Programme de soutien aux organismes de recherche et d’innovation to A.F. and by the Fondation du CHUM. This work was also supported by a CIHR foundation grant #352417, by a CIHR operating Pandemic and Health Emergencies Research grant #177958, to A.F., and by an Exceptional Fund COVID-19 from the Canada Foundation for Innovation (CFI) #41027 to D.E.K. and A.F. This work was also supported by a FRQS Pandemic Initiatives COVID-19 grant #308941 to M.J.H and in-kind contribution from Canadian Donation and Transplantation Research Program (CDTRP). Work on variants presented was also supported by the Sentinelle COVID Quebec network led by the LSPQ in collaboration with Fonds de Recherche du Québec Santé (FRQS) to A.F. A.F. is the recipient of Canada Research Chair on Retroviral Entry no. RCHS0235 950-232424. M.-J. H. holds the Shire Chair in Nephrology, Transplantation and Renal Regeneration of Université de Montréal. C.T holds the Pfizer/Université de Montréal Chair on HIV translational research. V.M.L. is supported by a FRQS Junior 1 salary award. D.E.K. is a FRQS Merit Research Scholar. G.B.B. is the recipient of a FRQS PhD fellowship. The funders had no role in study design, data collection and analysis, decision to publish, or preparation of the manuscript. We declare no competing interests.

## AUTHOR CONTRIBUTIONS

A.T., M.D., M.J.H. and A.F. conceived the study. A.T., G.B.B., S.Y.G., D.C., C.B., P.L., G.G.L. M.D., M.J.H. and A.F. performed, analyzed, and interpreted the experiments. A.T. and R.D. performed statistical analysis. G.B.B., S.Y.G., G.G.L., G.G., J.T., M.D., M.J.H. and A.F. contributed unique reagents. N.R., Z.K. C.T., D.E.K., M.J.H. and V.M.-L. collected and provided clinical samples. D.E.K., M.C., R.B., M.D., M.J.H. and A.F provided scientific input. A.T., and A.F. wrote the manuscript with inputs from others. Every author has read, edited, and approved the final manuscript.

## STAR METHODS

### RESOURCE AVAILABILITY

#### Lead contact

Further information and requests for resources and reagents should be directed to and will be fulfilled by the lead contact, Andrés Finzi.

#### Materials availability

All unique reagents generated during this study are available from the Lead contact without restriction.

#### Data and code availability

- All data reported in this paper will be shared by the lead contact upon request.
- This paper does not report original code.
- Any additional information required to reanalyze the data reported in this paper is available from the lead contact upon request.

## EXPERIMENTAL MODEL AND SUBJECT DETAILS

### Ethics Statement

All work was conducted in accordance with the Declaration of Helsinki in terms of informed consent and approval by an appropriate institutional board. Blood samples were obtained from donors who consented to participate in this research project at CHUM (19.381 and 21.001). Plasmas were isolated by centrifugation and Ficoll gradient, and samples stored at -80°C until use.

### Human subjects

The study was conducted in 20 SARS-CoV-2 naïve vaccinated HCW (8 males and 12 females; age range: 33-64 years) and 64 SARS-CoV-2 vaccinated SOTR (39 males and 25 females; age range: 21-82 years). All this information is summarized in table 1. For SARS-CoV-2 naïve vaccinated HCW cohort, no specific criteria such as number of patients (sample size), gender, clinical or demographic were used for inclusion, beyond no detection of Abs recognizing the N protein. For SARS-CoV-2 vaccinated SOTR, greater than 1-month post-transplant at the time of enrolment was used for inclusion.

### Plasma and antibodies

Plasma from SOTR and HCW were collected, heat-inactivated for 1 hour at 56°C and stored at - 80°C until ready to use in subsequent experiments. Plasma from uninfected donors collected before the pandemic were used as negative controls and used to calculate the seropositivity threshold in our ELISA, ADCC and flow cytometry assays (see below). The RBD-specific monoclonal antibody CR3022 was used as a positive control in our ELISA, and the CV3-25 antibody in flow cytometry assays and were previously described (Anand et al., 2020; Beaudoin-Bussières et al., 2020; Jennewein et al., 2021; Meulen et al., 2006; Prévost et al., 2020). Horseradish peroxidase (HRP)-conjugated Abs able to recognize the Fc region of human IgG (Invitrogen) were used as secondary Abs in ELISA experiments. Alexa Fluor-647-conjugated goat anti-human Abs able to detect all Ig isotypes (anti-human IgM+IgG+IgA; Jackson ImmunoResearch Laboratories) were used as secondary Abs to detect plasma binding in flow cytometry experiments.

### Cell lines

293T human embryonic kidney cells (obtained from ATCC) were maintained at 37°C under 5% CO_2_ in Dulbecco’s modified Eagle’s medium (DMEM) (Wisent) containing 5% fetal bovine serum (FBS) (VWR) and 100 μg/ml of penicillin-streptomycin (Wisent). CEM.NKr CCR5+ cells (NIH AIDS reagent program) and CEM.NKr.Spike cells were maintained at 37°C under 5% CO_2_ in Roswell Park Memorial Institute (RPMI) 1640 medium (Gibco) containing 10% FBS and 100 μg/ml of penicillin-streptomycin. 293T-ACE2 cell line was previously reported (Prévost et al., 2020). CEM.NKr CCR5+ cells stably expressing the SARS-CoV-2 S glycoprotein were previously reported (Anand et al., 2021; Beaudoin-Bussières et al., 2021).

## METHOD DETAILS

### Plasmids

The HCoV-HKU1 S expressing plasmid was purchased from Sino Biological. The plasmids encoding the different SARS-CoV-2 Spike variants (D614G, Delta and Omicron) were previously described (Beaudoin-Bussières et al., 2020; Chatterjee et al., 2022; Gong et al., 2021; Tauzin et al., 2021, 2022c).

### Protein expression and purification

FreeStyle 293F cells (Invitrogen) were grown in FreeStyle 293F medium (Invitrogen) to a density of 1 × 10^6^ cells/mL at 37°C with 8 % CO_2_ with regular agitation (150 rpm). Cells were transfected with a plasmid coding for SARS-CoV-2 S RBD (Beaudoin-Bussières et al., 2020) using ExpiFectamine 293 transfection reagent, as directed by the manufacturer (Invitrogen). One week later, cells were pelleted and discarded. Supernatants were filtered using a 0.22 μm filter (Thermo Fisher Scientific). The recombinant RBD proteins were purified by nickel affinity columns, as directed by the manufacturer (Invitrogen). The RBD preparations were dialyzed against phosphate-buffered saline (PBS) and stored in aliquots at -80°C until further use. To assess purity, recombinant proteins were loaded on SDS-PAGE gels and stained with Coomassie Blue.

### Enzyme-Linked Immunosorbent Assay (ELISA) and RBD avidity index

The SARS-CoV-2 RBD ELISA assay used was previously described (Beaudoin-Bussières et al., 2020; Prévost et al., 2020). Briefly, recombinant SARS-CoV-2 S RBD proteins (2.5 μg/ml) were prepared in PBS and were adsorbed to plates (MaxiSorp Nunc) overnight at 4°C. Coated wells were subsequently blocked with blocking buffer (Tris-buffered saline [TBS] containing 0.1% Tween20 and 2% BSA) for 1h at room temperature. Wells were then washed four times with washing buffer (Tris-buffered saline [TBS] containing 0.1% Tween20). CR3022 mAb (50 ng/ml) or a 1/250 dilution of plasma were prepared in a diluted solution of blocking buffer (0.1 % BSA) and incubated with the RBD-coated wells for 90 minutes at room temperature. Plates were washed four times with washing buffer followed by incubation with secondary Abs (diluted in a solution of blocking buffer (0.4% BSA)) for 1h at room temperature, followed by four washes. To calculate the RBD-avidity index, we performed in parallel a stringent ELISA, where the plates were washed with a chaotropic agent, 8M of urea, added of the washing buffer. This assay was previously described (Tauzin et al., 2022b). HRP enzyme activity was determined after the addition of a 1:1 mix of Western Lightning oxidizing and luminol reagents (Perkin Elmer Life Sciences). Light emission was measured with a LB942 TriStar luminometer (Berthold Technologies). Signal obtained with BSA was subtracted for each plasma and was then normalized to the signal obtained with CR3022 present in each plate. The seropositivity threshold was established using the following formula: mean of pre-pandemic SARS-CoV-2 negative plasma + (3 standard deviation of the mean of pre-pandemic SARS-CoV-2 negative plasma).

### Cell surface staining and flow cytometry analysis

293T cells were co-transfected with a GFP expressor (pIRES2-GFP, Clontech) in combination with plasmid encoding the full-length S of SARS-CoV-2 variants (D614G, Delta or Omicron) or the HCoV-HKU1 S. 48h post-transfection, S-expressing cells were stained with the CV3-25 Ab (Jennewein et al., 2021) or plasma (1/250 dilution). Alexa Fluor-647-conjugated goat anti-human IgM+IgG+IgA Abs (1/800 dilution) were used as secondary Abs. The percentage of transfected cells (GFP+ cells) was determined by gating the living cell population based on viability dye staining (Aqua Vivid, Invitrogen). Samples were acquired on a LSRII cytometer (BD Biosciences), and data analysis was performed using FlowJo v10.7.1 (Tree Star). The seropositivity threshold was established using the following formula: (mean of pre-pandemic SARS-CoV-2 negative plasma + (3 standard deviation of the mean of pre-pandemic SARS-CoV-2 negative plasma). The conformational-independent S2-targeting mAb CV3-25 was used to normalize Spike expression and was shown to effectively recognize all SARS-CoV-2 Spike variants (Li et al., 2022).

### ADCC assay

This assay was previously described (Anand et al., 2021; Beaudoin-Bussières et al., 2021). For evaluation of anti-SARS-CoV-2 ADCC, parental CEM.NKr CCR5+ cells were mixed at a 1:1 ratio with CEM.NKr cells stably expressing a GFP-tagged full length SARS-CoV-2 Spike (CEM.NKr.SARS-CoV-2.Spike cells). These cells were stained for viability (AquaVivid; Thermo Fisher Scientific, Waltham, MA, USA) and cellular dyes (cell proliferation dye eFluor670; Thermo Fisher Scientific) to be used as target cells. Overnight rested PBMCs were stained with another cellular marker (cell proliferation dye eFluor450; Thermo Fisher Scientific) and used as effector cells. Stained target and effector cells were mixed at a ratio of 1:10 in 96-well V-bottom plates. Plasma (1/500 dilution) were added to the appropriate wells. The plates were subsequently centrifuged for 1 min at 300g, and incubated at 37°C, 5% CO_2_ for 5 hours before being fixed in a 2% PBS-formaldehyde solution. ADCC activity was calculated using the formula: [(% of GFP+ cells in Targets plus Effectors)-(% of GFP+ cells in Targets plus Effectors plus plasma/antibody)]/(% of GFP+ cells in Targets) × 100 by gating on transduced live target cells. All samples were acquired on an LSRII cytometer (BD Biosciences) and data analysis was performed using FlowJo v10.7.1 (Tree Star). The specificity threshold was established using the following formula: (mean of pre-pandemic SARS-CoV-2 negative plasma + (3 standard deviation of the mean of pre-pandemic SARS-CoV-2 negative plasma).

### Virus neutralization assay

To produce the pseudoviruses, 293T cells were transfected with the lentiviral vector pNL4.3 R-E-Luc (NIH AIDS Reagent Program) and a plasmid encoding for the indicated S glycoprotein (D614G, Delta or Omicron) at a ratio of 10:1. Two days post-transfection, cell supernatants were harvested and stored at -80°C until use. For the neutralization assay, 293T-ACE2 target cells were seeded at a density of 1×10^4^ cells/well in 96-well luminometer-compatible tissue culture plates (Perkin Elmer) 24h before infection. Pseudoviral particles were incubated with several plasma dilutions (1/50; 1/250; 1/1250; 1/6250; 1/31250) for 1h at 37°C and were then added to the target cells followed by incubation for 48h at 37°C. Then, cells were lysed by the addition of 30 μL of passive lysis buffer (Promega) followed by one freeze-thaw cycle. An LB942 TriStar luminometer (Berthold Technologies) was used to measure the luciferase activity of each well after the addition of 100 μL of luciferin buffer (15mM MgSO_4_, 15mM KPO_4_ [pH 7.8], 1mM ATP, and 1mM dithiothreitol) and 50 μL of 1mM d-luciferin potassium salt (Prolume). The neutralization half-maximal inhibitory dilution (ID_50_) represents the plasma dilution to inhibit 50% of the infection of 293T-ACE2 cells by pseudoviruses.

## QUANTIFICATION AND STATISTICAL ANALYSIS

### Statistical analysis

Symbols represent biologically independent samples from SOTR and HCW. Lines connect data from the same donor. Statistics were analyzed using GraphPad Prism version 8.0.1 (GraphPad, San Diego, CA). Every dataset was tested for statistical normality and this information was used to apply the appropriate (parametric or nonparametric) statistical test. Differences in responses for the same donor after the second and third dose of mRNA vaccine were performed using Mann-Whitney tests. Differences in responses between HCW and SOTR at D2 or D3 were measured by Kruskal-Wallis tests. P values < 0.05 were considered significant; significance values are indicated as * p < 0.05, ** p < 0.01, *** p < 0.001, **** p < 0.0001. Spearman’s R correlation coefficient was applied for correlations. Statistical tests were two-sided and p < 0.05 was considered significant.

### Software scripts and visualization

Edge bundling graphs were generated in undirected mode in R and RStudio using ggraph, igraph, tidyverse, and RColorBrewer packages (R Core Team, 2014). Edges are only shown if p < 0.05, and nodes are sized according to the connecting edges’ r values. Nodes are color-coded according to groups of variables.

## SUPPLEMENTAL INFORMATION

Supplemental information can be found online at …

---

**Figure S1 :**
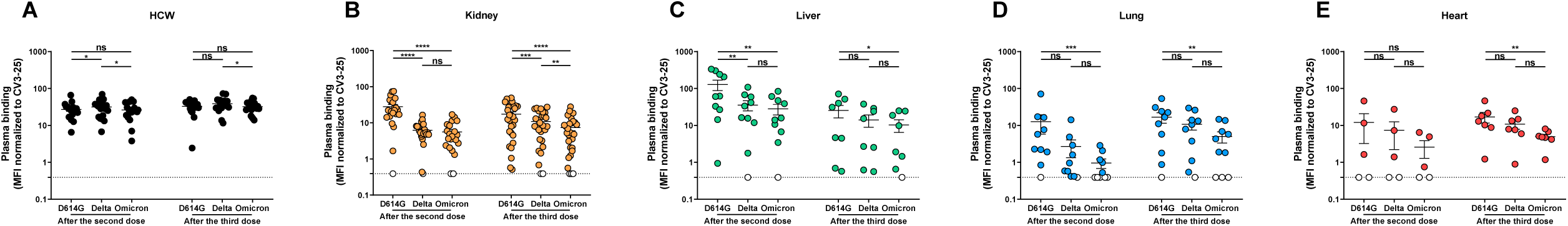
Recognition of SARS-CoV-2 Spike variants by vaccinated solid organ transplant recipients, Related to Figure 2. 293T cells were transfected with the indicated full-length S from different SARS-CoV-2 variants and stained with the CV3-25 Ab or with plasma from HCW (**A**) or transplant recipients (**B-E**) collected after the second and third doses of mRNA vaccine. The values represent the MFI normalized by the CV3-25 Ab. Symbols represent biologically independent samples from transplant recipients and HCW. Undetectable measures are represented as white symbols, and limits of detection are plotted. Error bars indicate means ± SEM. (* p < 0.05; ** p < 0.01; *** p < 0.001; **** p < 0.0001; ns, non-significant).

**Figure S2 :**
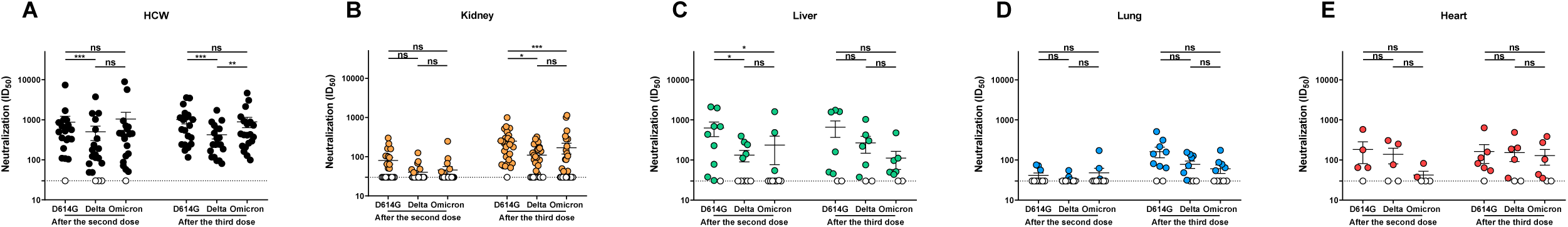
Neutralization of SARS-CoV-2 Spike variants by vaccinated solid organ transplant recipients, Related to Figure 3. Neutralizing activity was measured by incubating pseudoviruses bearing SARS-CoV-2 S glycoproteins with serial dilutions of plasma from HCW (**A**) and transplant recipients (**B-E**) collected after the second and third doses of mRNA vaccine for 1 h at 37°C before infecting 293T-ACE2 cells. Neutralization half maximal inhibitory serum dilution (ID_50_) values were determined using a normalized non-linear regression using GraphPad Prism software. Symbols represent biologically independent samples from transplant recipients and HCW. Undetectable measures are represented as white symbols, and limits of detection are plotted. Error bars indicate means ± SEM. (* p < 0.05; ** p < 0.01; *** p < 0.001; **** p < 0.0001; ns, non-significant).

